# Ultra-fast sample-to-sequencing workflow for clinical diagnostics using micropillars

**DOI:** 10.64898/2026.01.29.26345156

**Authors:** Adam J Bisogni, Irem Bastuzel, Mohamed Rashed, Joy Goffena, Sophie HR Storz, Zachary B. Anderson, Min Seon Park, Trent Prall, Miranda PG Zalusky, Erin E. Crotty, Bonnie Cole, Jeffrey Stevens, David M Lin, Harvey Tian, Danny E. Miller

**Affiliations:** Inso Biosciences, Ithaca, NY, 14853; Division of Genetic Medicine, Department of Pediatrics, University of Washington and Seattle Children’s Hospital, Seattle, WA, 98195; Department of Laboratory Medicine and Pathology, University of Washington, Seattle, WA, 98195; Division of Hematology, Oncology, Bone Marrow Transplant and Cellular Therapy, Department of Pediatrics, University of Washington and Seattle Children’s Hospital, Seattle, WA, 98195; Department of Laboratories, Seattle Children’s Hospital, Seattle, WA, 98105; Ben Towne Center for Childhood Cancer and Blood Disorders Research, Seattle Children’s Research Institute, Seattle, WA, 98101; Department of Biomedical Sciences, Cornell University, Ithaca, NY, 14853; Brotman Baty Institute for Precision Medicine, University of Washington, Seattle, WA, 98195

**Keywords:** Nanopore sequencing, long-read sequencing, DNA extraction, library preparation, microfluidics, point-of-care diagnostics

## Abstract

We present a streamlined, solid-phase workflow for Oxford Nanopore sequencing that integrates DNA extraction, purification, and library preparation within a single microfluidic cartridge. By eliminating tube transfers and performing all enzymatic steps directly on captured DNA, the method minimizes sample loss, reduces hands-on time, and simplifies library generation for long-read sequencing. Starting from volumes as small as a single drop of blood, this integrated approach produces high-quality sequencing libraries from cell lines, whole blood, and tissue. The workflow achieves robust recovery of high-molecular-weight DNA and high pore occupancy, enabling rapid, low-complexity sample preparation suitable for clinical, field, and decentralized sequencing applications.

## Main Text

There is substantial interest in the use of long-read sequencing (LRS) in both clinical and research settings as a tool to simplify the analysis of complex regions, directly detect sequence modifications, and reduce turnaround times.^1–3^ However, upstream sample preparation remains a significant bottleneck, especially in clinical workflows where DNA extraction and library preparation may take longer than sequencing itself to return an actionable result.^4,5^ This is especially true for rapid cancer diagnostics using methylation signatures, where near-real time diagnostics (<1 hour) is possible, but DNA extraction and library preparation may limit the time to result.^6,7^

Traditional workflows separate DNA extraction, purification, and library preparation into distinct phases, often requiring harsh mechanical homogenization or extensive pipetting that shears DNA, leads to sample loss, and may require substantial experience to perform well.^8^ To address these limitations, we developed an integrated “sample-to-sequence” workflow utilizing microfluidic micropillars that enables physical DNA capture and *in situ* library preparation **(Figure 1)**. By retaining the DNA on a high-surface-area solid support throughout lysis, washing, and sequencing library preparation, this approach minimizes mechanical shearing and eliminates the need for elution and re-binding steps common in magnetic bead-based protocols.^9–11^

**Figure 1.**
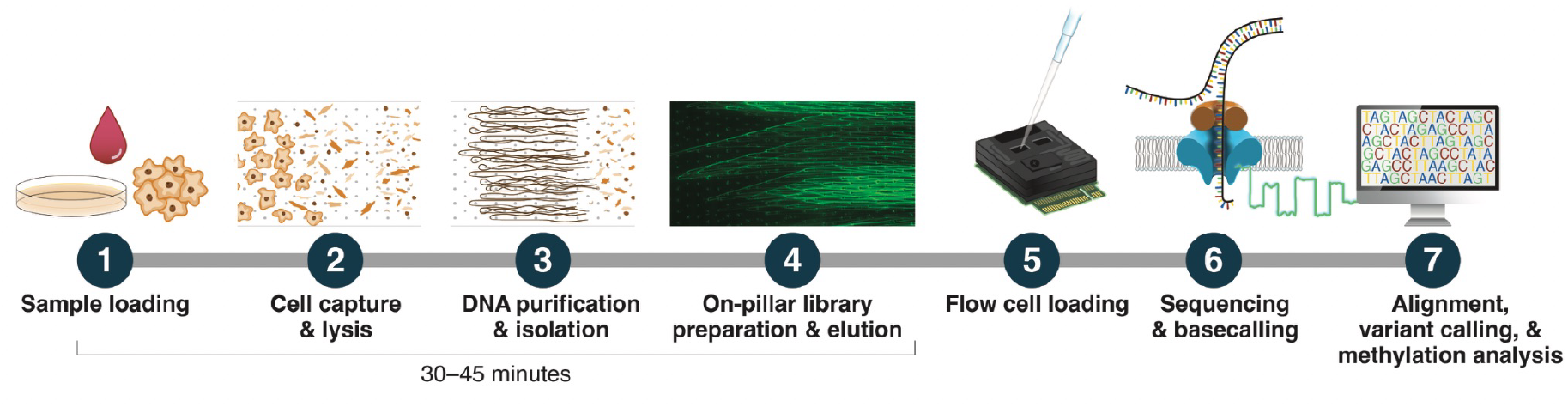
Overview of the integrated micropillar-based workflow for DNA isolation and library preparation for Nanopore sequencing. **(1)** Biological samples (whole blood, cultured cells, or dissociated tissue) are loaded directly onto a microfluidic cartridge containing a high-surface-area micropillar array. **(2)** Cells are physically captured within the micropillars and chemically lysed *in situ*. **(3)** Genomic DNA remains immobilized on the pillar surfaces during wash steps that remove inhibitors while preserving high-molecular-weight DNA. **(4)** ONT rapid library preparation reagents are introduced directly into the cartridge, enabling transposase-mediated adapter ligation on immobilized DNA (visualized here for demonstration purposes using fluorescence photomicrography with an intercalating dye), followed by elution from the pillars. **(5)** The completed sequencing library is loaded directly onto a Nanopore flow cell for sequencing **(6)** and analysis **(7)**. By eliminating tube transfers, centrifugation, and intermediate elution steps, this workflow minimizes DNA loss and mechanical shearing while substantially reducing hands-on time.

To validate this pillar-based approach for LRS sample preparation, we performed two sequencing runs using DNA isolated directly from deidentified whole blood (M3791 and M3792). Approximately 20 μL of whole blood was loaded directly onto the microfluidic cartridge where cells were captured and lysed on the pillars and DNA was extracted *in situ*. Immobilized DNA underwent rapid wash steps to remove contaminants before rapid transposase-based Oxford Nanopore (ONT) library preparation reagents were added directly to the cartridge, followed by elution of the completed library. Libraries were loaded onto used ONT PromethION R10.4.1 flow cells and sequenced for 21 hours. For each run, approximately 25 Gb of data was generated from 6.3 million reads with an average read length of 8–9 kbp giving a total aligned coverage of 7.1–7.7× **(Figure 2A, Table 1)**. Pore occupancy after 1 hour of sequencing was near 75%, excluding adapter-only reads (adapters that did not bind a DNA molecule for sequencing), suggesting the library preparation was of high-quality **(Figure 2B)**.

**Table 1.** Per-sample source, input amount, sequencing method, and sequencing results.

| Sample | Source | Input | Method | Starting Pores | Runtime (hours) | Gb | Total Reads (millions) | N50 (genome-wide) | N50 (target region) | Total Coverage (x) | Target Coverage (x) | Adaptive Sampling Enrichment (x) |
| --- | --- | --- | --- | --- | --- | --- | --- | --- | --- | --- | --- | --- |
| M3791 | WBCs | 20 ul whole blood | WGS | 4213 | 21.3 | 24 | 6.2 | 9,059 | - | 7.14 | - | - |
| M3792 | WBCs | 20 ul whole blood | WGS | 4233 | 21.3 | 25 | 6.4 | 8,560 | - | 7.68 | - | - |
| M3781 | LCL | 75,000 cells | AS | 6356 | 18.6 | 19 | 29.7 | 575 | 5,708 | 4.39 | 15.7 | 3.6 |
| M3782 | LCL | 75,000 cells | AS | 7198 | 18.6 | 23 | 34.0 | 577 | 6,312 | 5.69 | 22.6 | 4.0 |
| M3810 | Mouse cortex | 75,000 cells | AS | 7425 | 72 | 16 | 27.2 | 493 | 5,536 | 3.9 | 15.7 | 4.0 |
| M3983 | Pediatric brain tumor | 100 µL of dissociation mix | AS | 5423 | 19 | 4.6 | 10.1 | 457 | 1,056 | 0.78 | 1.41 | 1.8 |
| M3939 | Finger Prick | 20 ul whole blood | AS | 6525 | 72 | 12 | 15.5 | 1,425 | 20,147 | 4.5 | 26.2 | 5.8 |

**Figure 2.**
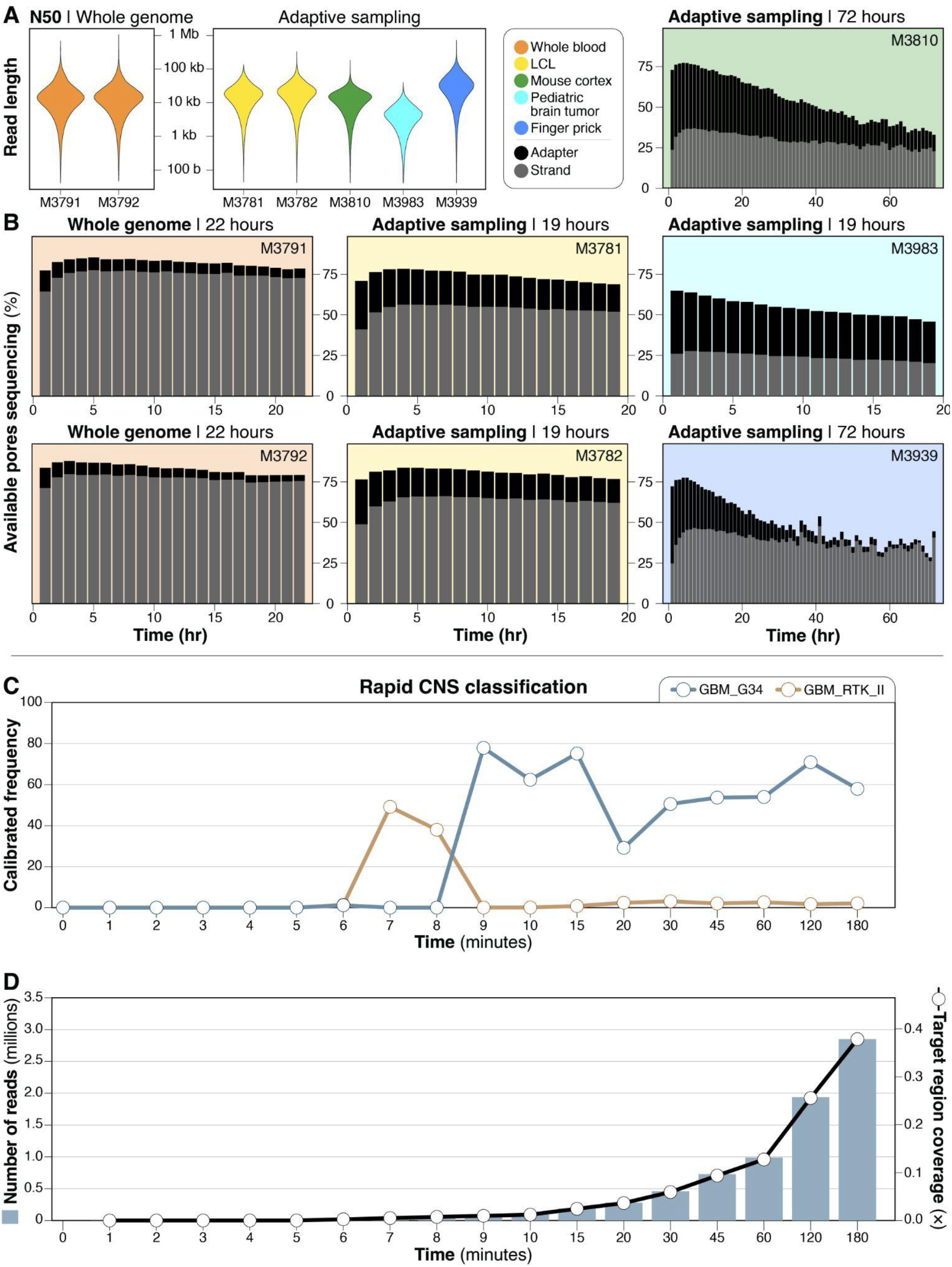
Sequencing performance of on-pillar library preparation across sample types and real-time tumor classification results. **(A)** Violin plots showing sequencing read length distributions (N50) for libraries generated from whole blood, cultured lymphoblastoid cells, tissue-derived samples, and blood collected via fingerstick. **(B)** Pore occupancy over sequencing time, separating adapter from adapter-only reads, illustrating sustained sequencing activity indicative of efficient library preparation and contaminant removal. **(C)** Retrospective real-time DNA methylation-based classification of a pediatric brain tumor sample (M3983), showing accurate identification of the tumor within 10 minutes of sequencing as a Glioblastoma, H3 G34–mutant (GBM_G34) using all available reads. All classifications with a probability >20% that appeared at least twice within 180 minutes are shown. **(D)** Genome coverage and number of reads generated over time for the pediatric brain tumor sample (M3983) showing that accurate tumor classification was possible with fewer than 100,000 reads.

We next performed two sequencing runs using the GM18865 cell line from Coriell (M3781 and M3782). For each sample, approximately 75,000 cells were loaded onto the microfluidic cartridge, and DNA extraction and library preparation were performed as described above. The prepared libraries were then eluted, loaded onto PromethION flow cells, and ran for 19 hours using an adaptive sampling protocol that targeted regions of interest in central nervous system (CNS) tumors **(Table S1)**.^12^ Sequencing yielded approximately 20 Gb of data per flow cell, with an on-target read N50 of ∼6 kbp **(Figure 2A, Table 1)**. Each sample generated 100,000 reads after ∼15 minutes of sequencing—a key metric for genome-wide methylation classification.^6^ After one hour, pore occupancy approached 50% for both samples, excluding adapter-only reads, again indicating high library quality and successful adapter ligation **(Figure 2B)**. This performance matches or exceeds standard affinity-binding protocols while reducing the total workflow time from approximately 1 hour, including ∼10 pipetting steps and 4 centrifuge steps, to approximately 30–45 minutes, with no centrifuge or pipetting steps. Sustained pore activity over the sequencing run confirmed that the library was of high quality and that the micropillar wash steps effectively removed contaminants that could affect the quality of the run. Despite the short sequencing time, both runs generated an on-target coverage of 16–23x, which allowed for variant calling and assessment of concordance within the target regions with F1 scores of 0.99 for single nucleotide variants, and 0.92 for insertion/deletion variants **(Table S2)**.

Given the growing interest in developing real-time sequencing applications for rapid brain tumor diagnostics,^6,13^ we applied the workflow to fresh brain tissue samples. Approximately 1 mg of mouse brain tissue was dissociated into cells, and 75,000 cells were then processed on-chip, yielding 202 ng of sequencing-ready DNA (M3810). Despite the high lipid content of brain tissue, which may clog spin columns or impede bead migration, the micropillar architecture facilitated effective clearance of cellular debris and allowed for successful on-pillar library preparation. An adaptive sampling sequencing run targeting mouse homologs of the same CNS tumor panel used for the cell-derived DNA **(Table S3)** was conducted for 72 hours on a PromethION flow cell. Pore occupancy was around 25% over the course of the run, which yielded 16.4 Gb of data **(Table 1)**. Coverage of the target regions was 15x representing a 4x enrichment over background.

To demonstrate utility for rapid methylation classification, we applied the workflow to a flash frozen pediatric brain tumor sample (M3983). We performed on-chip DNA extraction and library preparation using homogenate prepared from approximately 25 mg of frozen tumor tissue. The library was loaded onto a PromethION flow cell and sequenced for ∼19 hours with adaptive sampling targeting the same CNS tumor panel as above. This run yielded 4.6 Gb of data, with limited enrichment of target regions over background, likely reflecting constraints imposed by sample quality during DNA isolation and library preparation, including reduced pore occupancy and shorter fragment sizes **(Figure 2B)**. Regardless, we simulated real-time methylation classification of all reads (both on- and off-target) and found that we were able to accurately classify the tumor after ∼10 minutes of sequencing **(Figure 2C)**. Considered with the fresh tissue result above, this demonstrates the feasibility of our approach for rapid, intraoperative, sample-to-answer diagnostics of CNS tumors.

Finally, we evaluated the ability of our method to isolate DNA from capillary blood collected via finger stick (M3939). Approximately 20 μL of blood was applied directly from a finger onto the micropillar device and prepped for sequencing. The resulting library yielded 513 ng of DNA, which was sequenced on a new R10.4.1 flow cell using adaptive sampling targeting the CNS tumor panel. After 72 hours, 12.0 Gb of data were generated from 15.5 million reads, with a target region read N50 of 20.1 kbp and target region coverage of 26.2x, representing a 5.8x enrichment over background. Pore occupancy was approximately 40% during the run. These results demonstrate that the micropillar workflow can generate sequencing-quality libraries from minimally invasive sample collection, a capability particularly relevant for point-of-care diagnostics, pediatric applications, and resource-limited settings where phlebotomy infrastructure may be unavailable.^14,15^

In conclusion, we describe a micropillar-based “sample-to-sequence” workflow that demonstrates a unified solution for processing cells from culture, whole blood, and solid tissue with minimal manual intervention. By integrating extraction, purification, and library preparation into the micropillar architecture, the method preserves DNA integrity, reduces hands-on time, and minimizes the equipment footprint required for rapid LRS. Across blood, cultured cells, tissue inputs, and minimally invasive fingerstick samples this workflow enables rapid generation of sequencing-ready libraries compatible with real-time genomic and epigenomic analysis. These features make our approach well-suited for clinical diagnostics, rapid genomic screening where speed is critical, and field-based genomics applications.^14,16^

## METHODS

### Ethics approval

The institutional review board (IRB) of the University of Washington gave ethical approval for this work. The IRB of Seattle Children’s Hospital gave ethical approval for this work. Individual M3939 was enrolled in a study approved by the University of Washington IRB (Genomic Discovery Initiative, STUDY00020161), and written informed consent was obtained and archived by the communicating author. Individual M3983 was enrolled in a banking and biology study approved by the Seattle Children’s Hospital IRB (#14449). All other samples were deidentified prior to receipt and were not considered human subjects research.

### Sample collection and preparation

The lymphoblastoid cell line GM18865 was obtained from the Coriell Institute for Medical Research (Camden, NJ). Cells were maintained at a density of 200,000 to 1,000,000 cells per mL in RPMI 1640 supplemented with 2 mM L-glutamine and 15% fetal bovine serum and cultured at 37°C with 5% CO_2._ Prior to use, cells were pelleted by centrifugation at 100 × g for 10 minutes and resuspended in complete media to achieve the desired concentration. Anonymized adult human blood was obtained from Innovative Research (Novi, MI) and collected in K2 EDTA vacutainer tubes. For fingerstick whole blood collection, a sterile lancet was used to puncture the fingertip, and capillary blood was collected directly onto the micropillar device.

Mouse brain tissue was collected fresh and processed immediately using a Papain Dissociation System (LK003150, Worthington Biochemical Corporation, Lakewood, NJ). After dissociation the sample was loaded into the microfluidic cartridge for processing. Frozen tumor tissue was obtained from the Seattle Children’s Tumor Bank. To dissociate the tissue, a modified version of the QIAamp Fast DNA Tissue Kit (QIAGEN, Cat no. 51404) protocol was used. Briefly, approximately 25 mg of tissue was transferred to a bead-beating tube containing 265 µL of the tissue disruption buffer mix, as per the manufacturers’ instructions. Mechanical disruption was performed using a TissueLyser LT (QIAGEN) with two sequential bead-beating steps: 45 Hz for 60 seconds followed by 30 Hz for 45 seconds. The sample was then incubated on a heat block at 56°C for 10 minutes, with manual vortexing for 5–10 seconds every 3 minutes. Following the incubation, samples were immediately placed on ice. This homogenate was then directly loaded into the microfluidic cartridge for processing.

### Micropillar device fabrication

The micropillar devices were fabricated using established soft-lithography techniques as previously described.^9–11^ Briefly, the pillar architecture consists of a high-surface-area array designed to capture cells and retain DNA during washing and enzymatic processing steps. The micropillar architecture was optimized to enable efficient DNA binding while permitting release of the final library upon elution.

### On-pillar DNA isolation and library preparation

Samples were loaded directly onto the micropillar device, where cells were captured within the pillar matrix under constant flow conditions. On-pillar lysis was performed using QIAGEN Buffer AL, which is compatible with the chaotropic lysis approach required for efficient DNA release. Following lysis, the micropillar array was washed with 1× PBS to remove cellular debris and potential sequencing inhibitors while retaining high-molecular-weight DNA bound to the pillar surface. Library preparation was performed on-chip, directly on the immobilized DNA using the ONT Rapid Sequencing Kit (SQK-RAD114). A reaction mixture containing transposase was loaded into the microfluidic cartridge applied to the pillars, enabling library preparation without prior elution. Following the enzymatic reaction, the library was eluted from the pillars and prepared for loading onto the flowcell.

### Sequencing

Samples were sequenced on a PromethION 24 instrument running MinKNOW version 25.03.7 using R10.4.1 flowcells. For each sample, ∼19 μL of the library was loaded and sequenced for 19–72 hours depending on the experimental objectives. Two samples (M3791, M3792) were sequenced using a whole-genome approach, and five samples (M3781, M3782, M3810, M3683, M3693) were sequenced using adaptive sampling to enrich regions of interest. For adaptive sampling of human samples, we used an in-house-developed tumor gene panel comprising 841 genes using GRCh38 genomic coordinates **(Table S1)**. The mouse brain tissue sample (M3810) was sequenced with a corresponding panel containing 860 mouse orthologs using mm39 genomic coordinates **(Table S3)**. Orthologous gene mappings were obtained from the Mouse Genome Informatics database.^17^

### Bioinformatics analysis

After sequencing, POD5 files were basecalled using Dorado (v1.1.1) with the super accuracy model and 5mCG_5hmCG modifications enabled. Basecalled reads were aligned to either GRCh38 (for human samples) or mm39 (for mouse samples) using Minimap2 (v2.28).^18^ Single nucleotide variants were called using Clair3 (v1.0.8)^18,19^, and structural variants were called using Sniffles2 (v2.3.3).^20^ Aligned BAM files were phased using Longphase (v1.7.3).^21^ SNP concordance between adaptive sampling runs of GM18865 was calculated using GenotypeConcordance from the Picard Toolkit (v3.1.1). Input VCFs were restricted to the gene coordinates targeted in adaptive sampling and masked to highly reproducible regions of GRCh38. These regions contain reproducible variants from seven publicly available datasets and exclude unusual regions identified by UCSC, the ENCODE Blacklist, and exclusions identified by the genome reference consortium as false duplications or contamination sequences.^22^ Additionally, variants within one base pair of a homopolymer greater than 5 base pairs in length were excluded from concordance calculations.

### Tumor classification

Aligned reads were separated into temporal bins based on read_id and the corresponding start_time output by MinKNOW during sequencing. Methylation levels were quantified using the EPI2ME labs wf-human-variation workflow (v2.7.3) with the --bam_min_coverage flag set to 0 to generate bedmethyl files for each temporal bin.^23^ Tumor classification for each bin was determined utilizing Rapid-CNS2.^24^ The resulting calibrated frequency scores, specific to each timepoint, were plotted using the matplotlib library.

## Data Availability

All data produced in the present study are available upon reasonable request to the authors.

## Acknowledgements

We thank Angela Miller for expert assistance with figure preparation and editing. Funding for this project was provided by the Pediatric Brain Tumor Research Fund (PBTRF). DEM is supported by the National Institutes of Health through the NIH Director’s Early Independence Award DP5OD033357.

## References

1. Logsdon, G. A., Vollger, M. R. & Eichler, E. E. Long-read human genome sequencing and its applications. Nat. Rev. Genet. 21, 597–614 (2020).

2. Mantere, T., Kersten, S. & Hoischen, A. Long-Read Sequencing Emerging in Medical Genetics. Front Genet 10, 426 (2019).

3. Wojcik, M. H. et al. Beyond the exome: What’s next in diagnostic testing for Mendelian conditions. Am J Hum Genet 110, 1229–1248 (2023).

4. Zalusky, M. P. et al. 3-hour genome sequencing and targeted analysis to rapidly assess genetic risk. Genet Med Open 2, (2024).

5. Gorzynski, J. E. et al. Ultrarapid Nanopore Genome Sequencing in a Critical Care Setting. N. Engl. J. Med. 386, 700–702 (2022).

6. Djirackor, L. et al. Intraoperative DNA methylation classification of brain tumors impacts neurosurgical strategy. Neurooncol Adv 3, vdab149 (2021).

7. Capper, D. et al. DNA methylation-based classification of central nervous system tumours. Nature 555, 469–474 (2018).

8. Pinzauti, D., Iannelli, F., Pozzi, G. & Santoro, F. DNA isolation methods for Nanopore sequencing of the genome. Microb Genom 8, (2022).

9. Benítez, J. J. et al. Microfluidic extraction, stretching and analysis of human chromosomal DNA from single cells. Lab Chip 12, 4848–4854 (2012).

10. Tian, H. C., Benitez, J. J. & Craighead, H. G. Single cell on-chip whole genome amplification via micropillar arrays for reduced amplification bias. PLoS One 13, e0191520 (2018).

11. Reinholt, S. J. & Craighead, H. G. Microfluidic Device for Aptamer-Based Cancer Cell Capture and Genetic Mutation Detection. Anal Chem 90, 2601–2608 (2018).

12. Payne, A. et al. Readfish enables targeted nanopore sequencing of gigabase-sized genomes. Nat. Biotechnol. 39, 442–450 (2021).

13. Vermeulen, C. et al. Ultra-fast deep-learned CNS tumour classification during surgery. Nature 622, 842–849 (2023).

14. Quick, J. et al. Real-time, portable genome sequencing for Ebola surveillance. Nature 530, 228–232 (2016).

15. Meredith, L. W. et al. Rapid implementation of SARS-CoV-2 sequencing to investigate cases of health-care associated COVID-19: a prospective genomic surveillance study. Lancet Infect Dis 20, 1263–1272 (2020).

16. Wang, Y., Zhao, Y., Bollas, A., Wang, Y. & Au, K. F. Nanopore sequencing technology, bioinformatics and applications. Nat Biotechnol 39, 1348–1365 (2021).

17. Blake, J. A. et al. Mouse Genome Database (MGD): Knowledgebase for mouse-human comparative biology. Nucleic Acids Res 49, D981–D987 (2021).

18. Li, H. Minimap2: pairwise alignment for nucleotide sequences. Bioinformatics 34, 3094–3100 (2018).

19. Zheng, Z. et al. Symphonizing pileup and full-alignment for deep learning-based long-read variant calling. Nature Computational Science 2, 797–803 (2022).

20. Smolka, M. et al. Detection of mosaic and population-level structural variants with Sniffles2. Nat Biotechnol 42, 1571–1580 (2024).

21. Lin, J.-H., Chen, L.-C., Yu, S.-C. & Huang, Y.-T. LongPhase: an ultra-fast chromosome-scale phasing algorithm for small and large variants. Bioinformatics (2022) doi:10.1093/bioinformatics/btac058.

22. Pan, B. et al. Assessing reproducibility of inherited variants detected with short-read whole genome sequencing. Genome Biol 23, 2 (2022).

23. Website. https://github.com/nanoporetech/wf-human-variation.

24. Patel, A. et al. Rapid-CNS: rapid comprehensive adaptive nanopore-sequencing of CNS tumors, a proof-of-concept study. Acta Neuropathol 143, 609–612 (2022).

